# Fully Automated Explainable Abdominal CT Contrast Media Phase Classification Using Organ Segmentation and Machine Learning

**DOI:** 10.1101/2023.12.16.23299369

**Authors:** Yazdan Salimi, Zahra Mansouri, Ghasem Hajianfar, Amirhossein Sanaat, Isaac Shiri, Habib Zaidi

## Abstract

**Purpose:** To detect contrast media injection phase from CT images by means of organ segmentation and deep learning.

**Materials and Methods:** A total number of 2509 CT images split into four subsets of non-contrast (class #0), arterial (class #1), venous (class #2), and delayed (class #3) after contrast media injection were collected from two CT scanners. Seven organs including the liver, spleen, heart, kidneys, lungs, urinary bladder, and aorta along with body contour masks were generated by pre-trained deep learning algorithms. Subsequently, five first-order statistical features including average, standard deviation, 10, 50, and 90 percentiles extracted from the above-mentioned masks were fed to machine learning models after feature selection and reduction to classify the CT images in one of four above mentioned classes. A ten-fold data split strategy was followed. The performance of our methodology was evaluated in terms of classification accuracy metrics.

**Results:** The best performance was achieved by Boruta feature selection and RF model with average area under the curve of more than 0.999 and accuracy of 0.9936 averaged over four classes and ten folds. Boruta feature selection selected all predictor features. The lowest classification was observed for class #2 (0.9888), which is already an excellent result. In the ten-fold strategy, only 33 cases from 2509 cases (∼1.4%) were misclassified.

**Conclusion:** We developed a fast, accurate, reliable, and explainable methodology to classify contrast media phases which may be useful in data curation and annotation in big online datasets or local datasets with non-standard or no series description.

**Key points:**

1. The lack of standard series description and information about contrast media phase limits the usability of medical CT data.
2. We developed a twostep deep learning/machine learning solution with excellent performance.
3. This fast, automated, reliable and explainable purposed pipeline can tag every CT images with using only image matrices.

## Introduction

Computed tomography (CT), as a versatile non-invasive imaging modality, plays a significant role in clinical diagnosis and follow-up of patients presenting with a wide range of pathologies. The administration of contrast media (mostly positive contrast media) improves the diagnostic value and accuracy of this imaging procedure by enhancing the contrast in tissues [1]. This enhanced contrast is useful in the differentiation of multiple pathologic situations reflecting improved discrimination between different tissue types [2]. The most common method of contrast media administration for contrast enhanced CT (CE-CT) is intravenous injection using radiopaque materials mostly containing iodine. For instance, the management of patients treated with selective internal radiation therapy (SIRT) for the treatment of liver malignancies requires CE-CT for the delineation of the tumoral tissues, perfused liver lobe and organs at risk towards personalized dosimetry [3].

Artificial intelligence (AI) and particularly deep learning has shown very promising performance in multiple tasks, including image segmentation [4-6], image generation [7; 8], dosimetry [9-11], and classification [12; 13]. However, the number of clean and reliable data available is still the bottle neck for generalizability and robustness of deep learning models. In this regard, data and model sharing between imaging centers may help to collect diverse training datasets. However, even for federated learning strategies, the data should be in the same format to be able to train a model among different centers [14]. At the same time, due to privacy issues the image data needs to be anonymized by removing private tags, which may include removing the injection phase information. We evaluated the number of online available datasets and came to the conclusion that most datasets in DICOM format did not have any straightforward information about the contrast media injection phase, while the image only data formats, such as NIFTI, do not include any related information. Besides, within the same hospital when using different scanners, the naming of study/series/contrast injection phase are commonly not consistent [15], even in the same department. The formal written language between different cities and imaging centers is another barrier in effective data sharing between centers even in a single country. These limitations and lack of trustable information about the contrast media administrative phases underscore the need for developing an easily accessible solution based on minimum shared data, which is the image matrix only.

Philbrick et al. [16] trained deep neural network classifiers to separate contrast enhanced phases in abdominal CTs for kidney pathologies indications. They attempted to explain what the network sees during training and inference by generating GradCam and saliency maps after each decision. They reported F1-scores between 0.781 and 0.999 for different contrast media phases. Zhou et al. [17] trained a deep neural network to separate 4 phases of liver CE-CT including non-contrast, arterial, venous, and delay. They used natural language processing to extract the ground truth data from DICOM tags and reported an average F1-score of 0.977 on a very large dataset. Tang et al. [18] used a generative adversarial network to classify the contrast phases in abdominal CE-CTs and achieved an accuracy of 0.93. They used 2D axial slices to train and evaluate the algorithm. Dao et al. [19] used random sampling and deep neural networks for automated classification of CE-CT images. They reported F1 score of 0.9209 on the internal test consisting of 358 scans using 2D slices as input to the network. The evaluation of their algorithm on external datasets reported F1 scores of 0.7679 and 0.8694 on two manually annotated online databases. Muhamedrahimov et al. [20] used CE-CT images and the time from injection as the ground truth to train a regression machine learning model to predict the time from injection. Then they used these times to classify images and reported an overall accuracy of 0.933 in classification. Rocha et al. [15] used machine learning to classify axial 2D slices from liver CE-CTs in four phases and reported an accuracy of more than 0.98. Reis et al. [21] independently adopted a strategy similar to the one used in our study and developed a machine learning method to classify contrast media phase based on few organs segmentation. They used online available datasets to train their network and reported F1 scores of 0.966, 0.789, 0.922, and 0.95 for four contrast phases consisting of non-contrast, arterial, venal, and delayed, respectively.

This study is an extension of previous preliminary work aimed to develop explainable, fast, and accurate methodology to classify the contrast media phase using imaging only data [22]. Inspired by the logic human follows to understand the injection phase, which includes checking the enhancement ratio and the HU in few organs, such as aorta and kidney, we attempted to use the very simple features extracted from the above mentioned regions and machine learning algorithms to classify CT images.

## Materials and Methods

This study included 2509 Thorax/Abdomen CT images acquired between 2011 and 2022 on scanners commercialized by two different vendors (Siemens Healthcare and Philips Medical Systems) in different imaging centers for evaluation of different liver pathologies, mainly for initial diagnosis and follow-up of liver malignancies, such as hepatocellular carcinoma (HCC). CT images were acquired in four phases, including non-enhanced (W/O) (class 0), arterial (class 1), venous (class 2), and delayed (class 3) acquisition with the same scan range covering the abdominal area or more. The pipeline of this study is showed in Figure 1. First, the technologists classified the images in one of the phases according to the DICOM series information headers. These classes for each 3D image were used as the ground truth for training and evaluation of our methodology. Meanwhile, seven organs including the liver, spleen, heart, kidneys, lungs, urinary bladder, and aorta plus body contour were segmented from CT images automatically using previously developed algorithm [23]. Then, five first-order features, including the average, standard deviation (SD), ten percentile, median, and 90 percentiles were extracted using basic Python software-based image processing. These five features for seven organs were used to train a machine learning algorithm to perform classification task in a ten-fold data split strategy. Then, the performance of the whole algorithm was evaluated in terms of F1 score, accuracy, specificity, sensitivity, precision, and area under the curve (AUC). Receiver operating characteristic (ROC) curves and confusion matrices were then generated for further evaluation.

**Figure 1.**
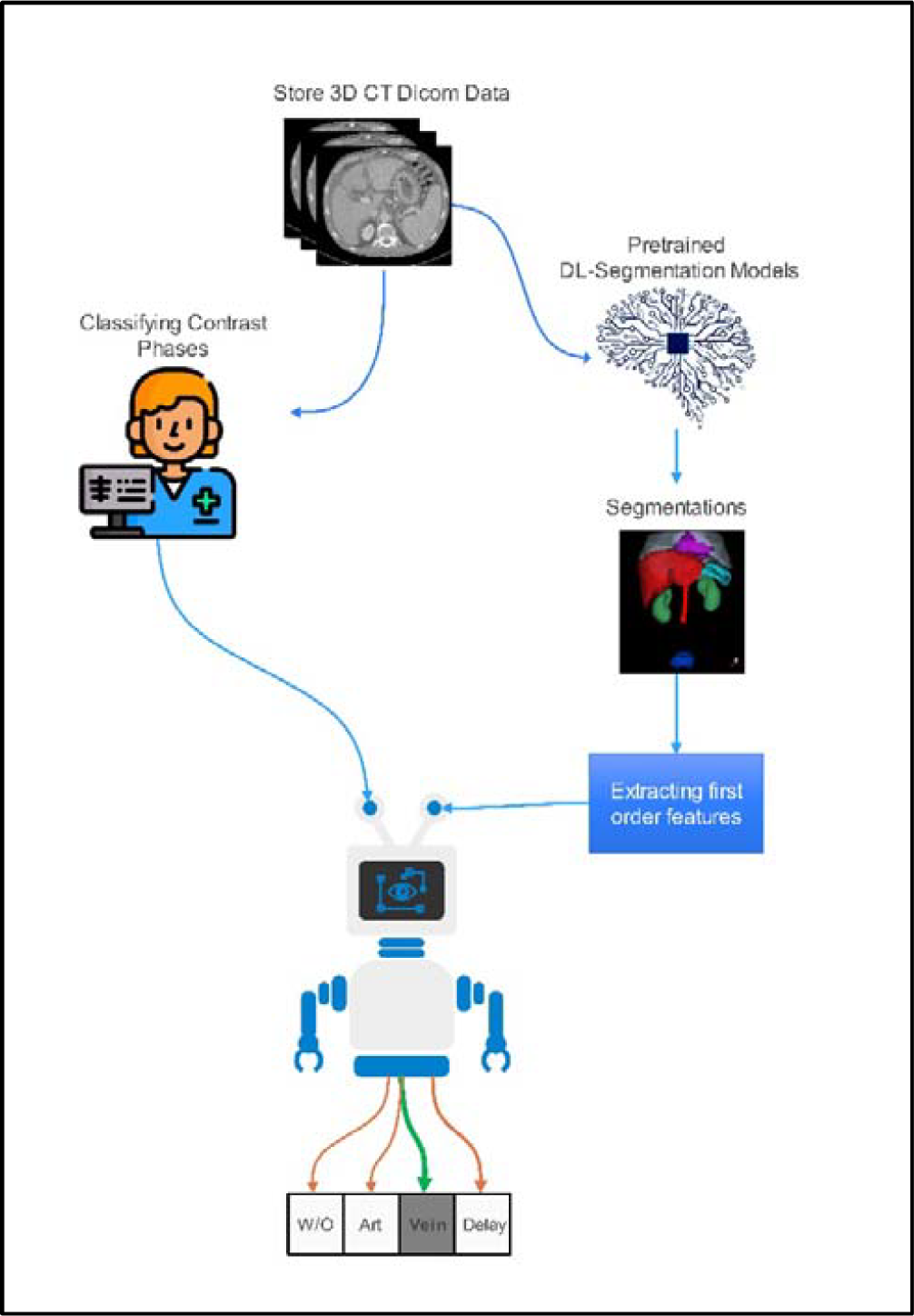
Flowchart of the steps followed in this study. The manual image labels were extracted according to the DICOM metadata.

### Feature selection and machine learning

Three feature selection methods, including Boruta [24], Maximum Relevance Minimum Redundancy (MRMR) [25], and Recursive Feature Elimination (RFE) [26] were used to reduce the number of features. Boruta and RFE are wrapper-based whereas MRMR is filter-based algorithm. In Boruta and RFE, the number of selected features is determined by specific criteria, while in MRMR, it is based on the top 15 features. Subsequently, 6 different classifiers, including K-nearest neighbors (KNN) [27], eXtreme Gradient Boosting (XGB) [28], Decision Tree (DT) [29], Support Vector Machine (SVM) [30], Naive Bayes (NB) [31], and Random Forest (RF) [32] were used to classify images. In this study, we used nested cross-validation with outer and inner validation. We used 10-fold cross-validation for outer validation. In each repetition of outer validation, feature selection methods were performed on 9 portions of data followed by training the classifier model. Hyperparameter optimization for each classifier was performed using inner 10-fold cross-validation scheme. The trained models with the best hyperparameter were tested on one portion of dataset. This procedure was repeated 10 times. Finally, among these 3 × 6 feature selection/models, the best model based on performance was selected. For model evaluation, we used confusion matrix, AUC, F1_score, sensitivity, specificity, precision, and accuracy.

### Segmentation model

A previously developed and validated ResUNET-based model was used for CT image segmentation [23]. First, the body contour was generated on CT images via image processing algorithms and then this contour was used to crop the images to foreground. Then, the trained models were used to generate segmentation masks through inference on every 2D axial CT slice. Organ-specific post-processing was performed to refine the network output by implementing prior knowledge about the position of organs in the human body. The Dice coefficients (in %) for this segmentation model reported on external dataset were 96.98 ± 1.62, 94.68 ± 9.28, 91.95 ± 6.13, 94.13 ± 5.05, 97.63 ± 1.18, 83.78 ± 17.97, 93.55 ± 2.22, for the liver, spleen, heart, kidneys, lungs, urinary bladder, aorta, respectively.

The five mentioned features including average, standard deviation (SD), ten percentile, median, and 90 percentiles were calculated inside the eight segmentation masks including the liver, spleen, heart, kidneys, lungs, urinary bladder, aorta, and body contour and recorded for the next step.

## Results

Table 1 summarizes the demographic information of the patients included in this study plus the acquisition/reconstruction CT parameters. Patients’ age was 65.23 ± 11.36 years, and the average tube current was 288.8989 ± 137.9301 mA. This was a balanced distribution of data between classes where there were 683, 714, 419, and 693 images in classes zero to three, respectively.

**Table 1.** Demographic and acquisition parameters of patients included in the study protocol.

| Age (years) | Date | Manufacturer | kVp | Pitch Factor | CTDI<sub>vol</sub> | Slice Thickness | Acquisition Diameter (mm) | Reconstruction Diameter (mm) | Weight (Kg) | Average Tube Current |
| --- | --- | --- | --- | --- | --- | --- | --- | --- | --- | --- |
| $65.23 \pm 11.36$ | 2011 to 2022 | Philips, Siemens | 70, 80, 90, 110, 130 | $0.90 \pm 0.25$ | $9.98 \pm 10.14$ | $2.35 \pm 1.68$ | $498 \pm 43$ | $414 \pm 84$ | $79.25 \pm 16.64$ | $288.89 \pm 137.93$ |

### Segmentation

Figure 2 and Figure 3 depict examples of visualization of segmented organs on axial images. The segmentation shows almost the same region in four phases. Please note the enhancement and differences in the segmented regions on Figure 2 and Figure 3. The urinary bladder was not included in all images. Hence, we excluded this organ as a predictor in our model i.e., six organs plus body contour (seven segmentation masks) were used to train the machine learning model. Figure 4 presents a 3D visualization of the segmented organs in different views. 3D rotating views showing the segmentation outcome are provided in supplementary material section.

**Figure 2.**
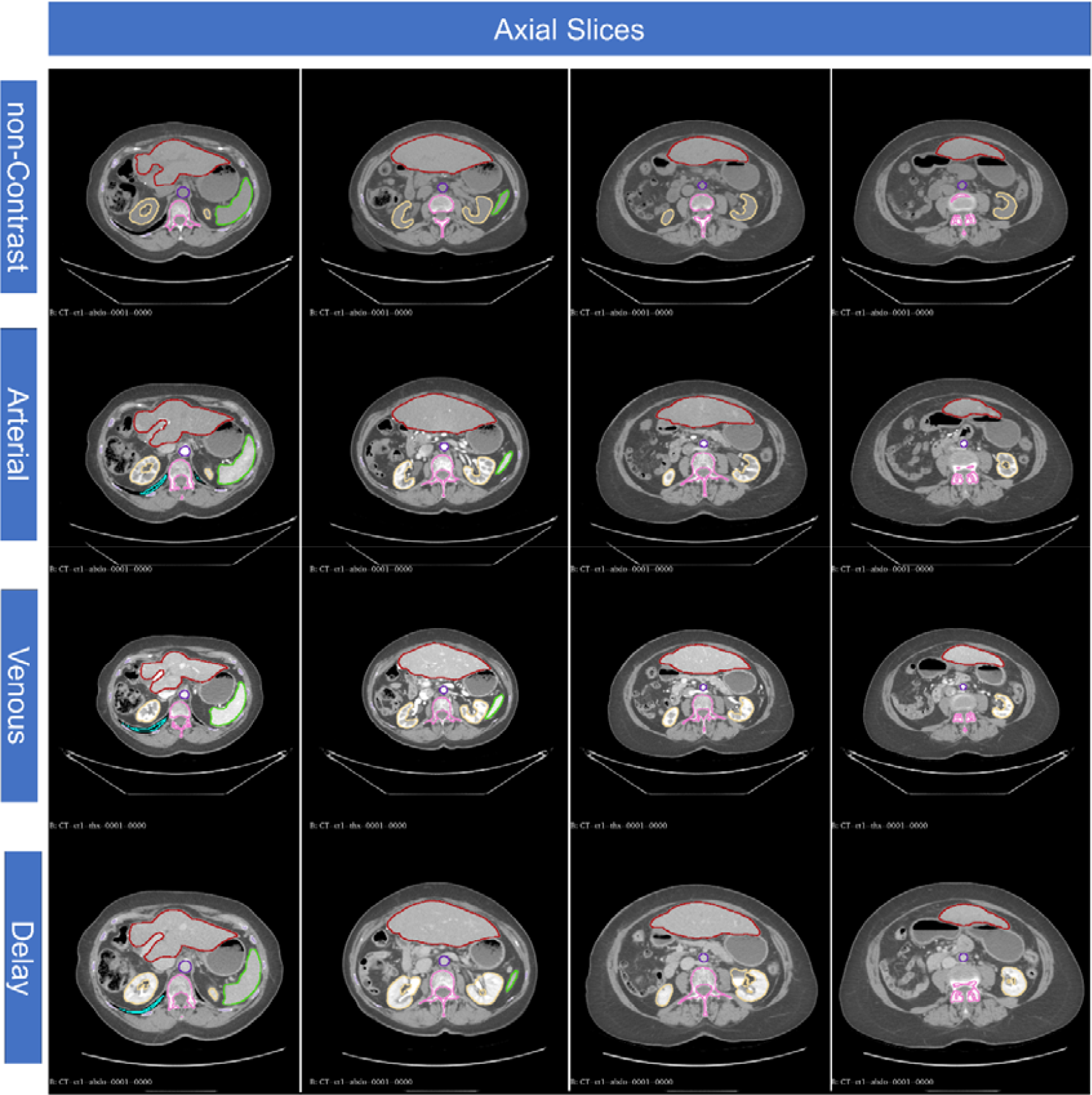
Axial CT slices of the same patient presenting with HCC in four phases including non-contrast (#0, top row), arterial (#1, second row), Venous (#2, third row) and delayed (#3, fourth row). The same line colors show the segmented organs. Please note the enhancement in the segmented regions. A window/level of 500/0 HU was used for all images to be comparable. Green: Spleen, Yellow: Kidneys, Dark Red: Liver, Light Blue: Lungs, Purple: Aorta, Pink: Vertebrae (was not used in the model).

**Figure 3.**
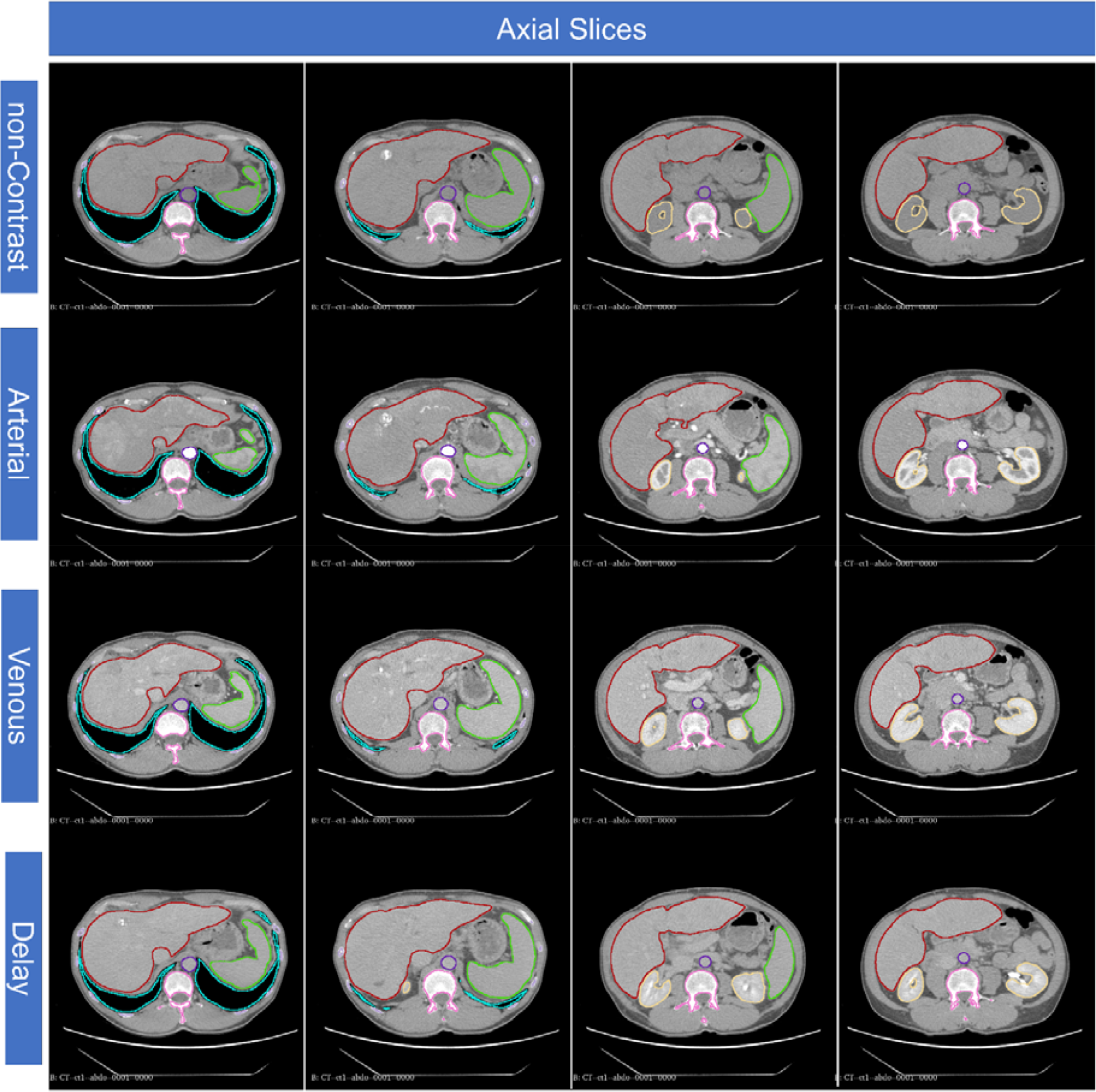
Axial CT slices for a patient referred for metastatic liver malignancies indication (same caption as Figure 2). The window/level is 500/0 HU and the difference in lung is not visible. Green: Spleen, Yellow: Kidneys, Dark Red: Liver, Light Blue: Lungs, Purple: Aorta, Pink: Vertebrae (was not used in the model).

**Figure 4.**
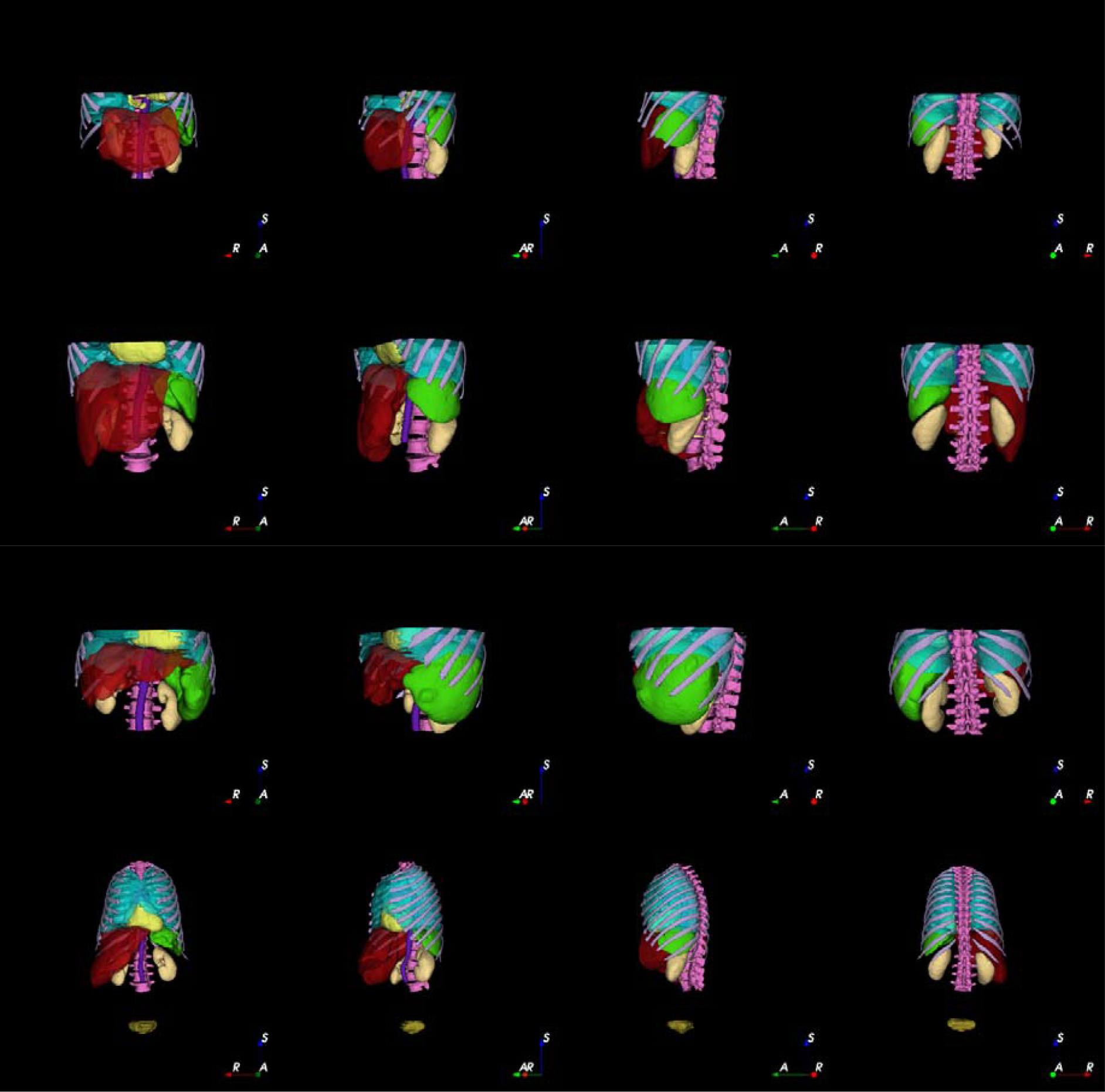
Examples of segmented organs displayed in 3D views. Yellow: Heart, Purple: Aorta, Brown: Urinary Bladder. Green: Spleen, Light yellow: Kidney, Dark Red: Liver, light blue: Lungs. The vertebrae and ribs were shown for better visualization. Each row belongs to a single patient and the columns represent 3D views from four different views from anterior to posterior.

### Selected model and feature selection

The best performance was achieved by Boruta feature selection and RF model. This feature selection method found all the expected important features, because the features were selected according to the specific task. The highest importance in Boruta feature selection was for aorta, cardiac, and spleen organs. The heatmap comparing the performance of different combinations of feature selection and models is presented in Supplementary figure 1.

### Classification results average for the best model/feature selection

Table 2 summarizes the results of the classification model calculated in ten-fold data split strategy. F1-score was 0.9842 in average for the four classes, while the highest score was for non-enhanced (0.9978) and the lowest was for venous phase (0.9667). An AUC higher than 0.99 was achieved for all four classes.

**Table 2.** The classification performance of the best model for each of the four classes.

| Class | F1_score | Sensitivity | Specificity | Precision | Accuracy | AUC |
| --- | --- | --- | --- | --- | --- | --- |
| **0 (W/O)** | 0.9978 | 0.9956 | 1.0000 | 1.0000 | 0.9988 | 0.9989 |
| **1 (Arterial)** | 0.9951 | 0.9916 | 0.9994 | 0.9986 | 0.9972 | 1.0000 |
| **2 (Vein)** | 0.9667 | 0.9690 | 0.9928 | 0.9644 | 0.9888 | 0.9983 |
| **3 (Delay)** | 0.9813 | 0.9856 | 0.9912 | 0.9771 | 0.9896 | 0.9991 |
| **Total Classes** | 0.9852 | 0.9854 | 0.9959 | 0.9850 | 0.9936 | 0.9991 |

Figure 5 represents the normalized confusion matrix for all data included in ten-fold strategy. The lowest performance was achieved for the venous phase once again.

**Figure 5.**
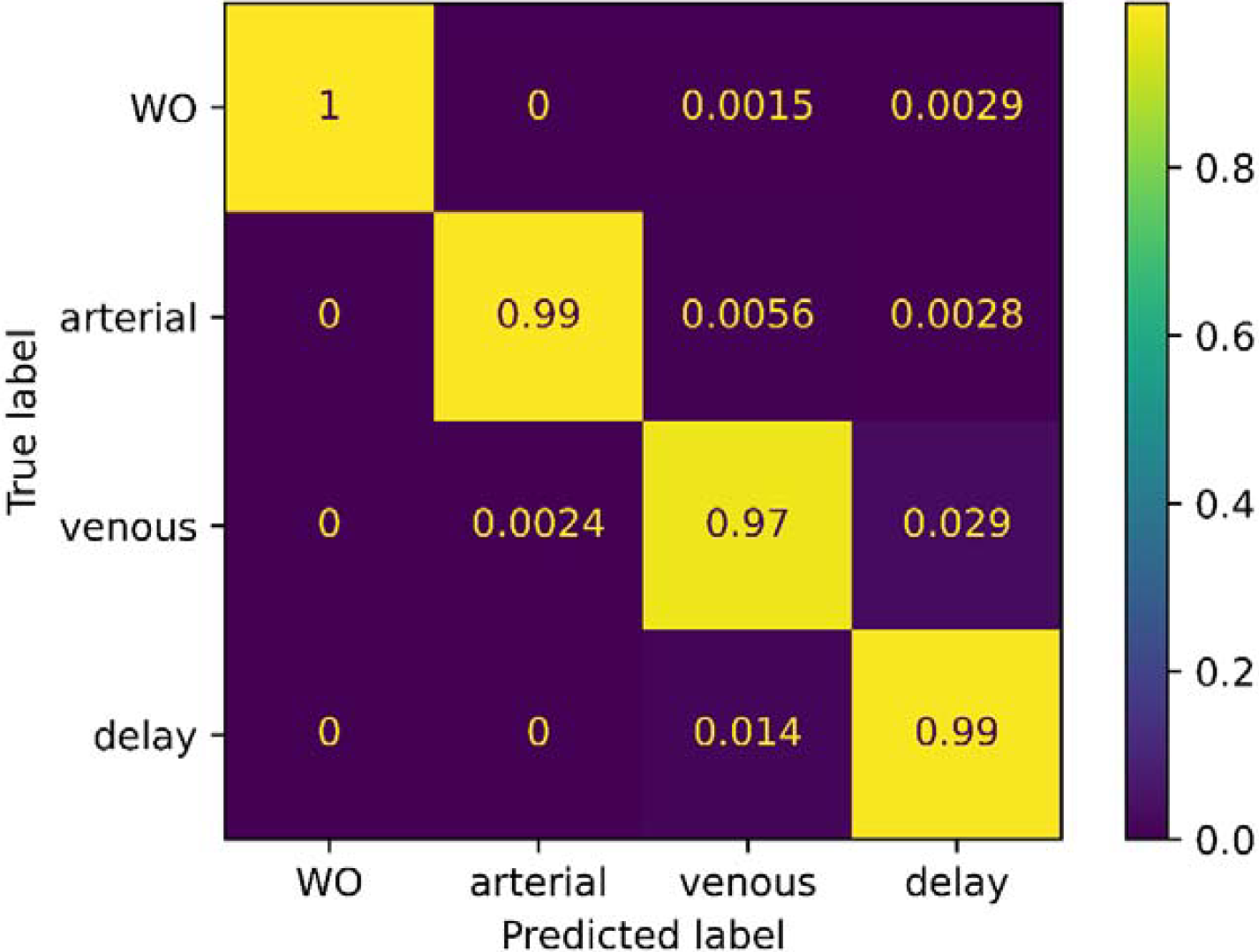
Normalized confusion matrix showing the excellent performance of the model in ten-fold strategy.

Figure 6 shows the ROC curves for all classes demonstrating excellent performance.

**Figure 6.**
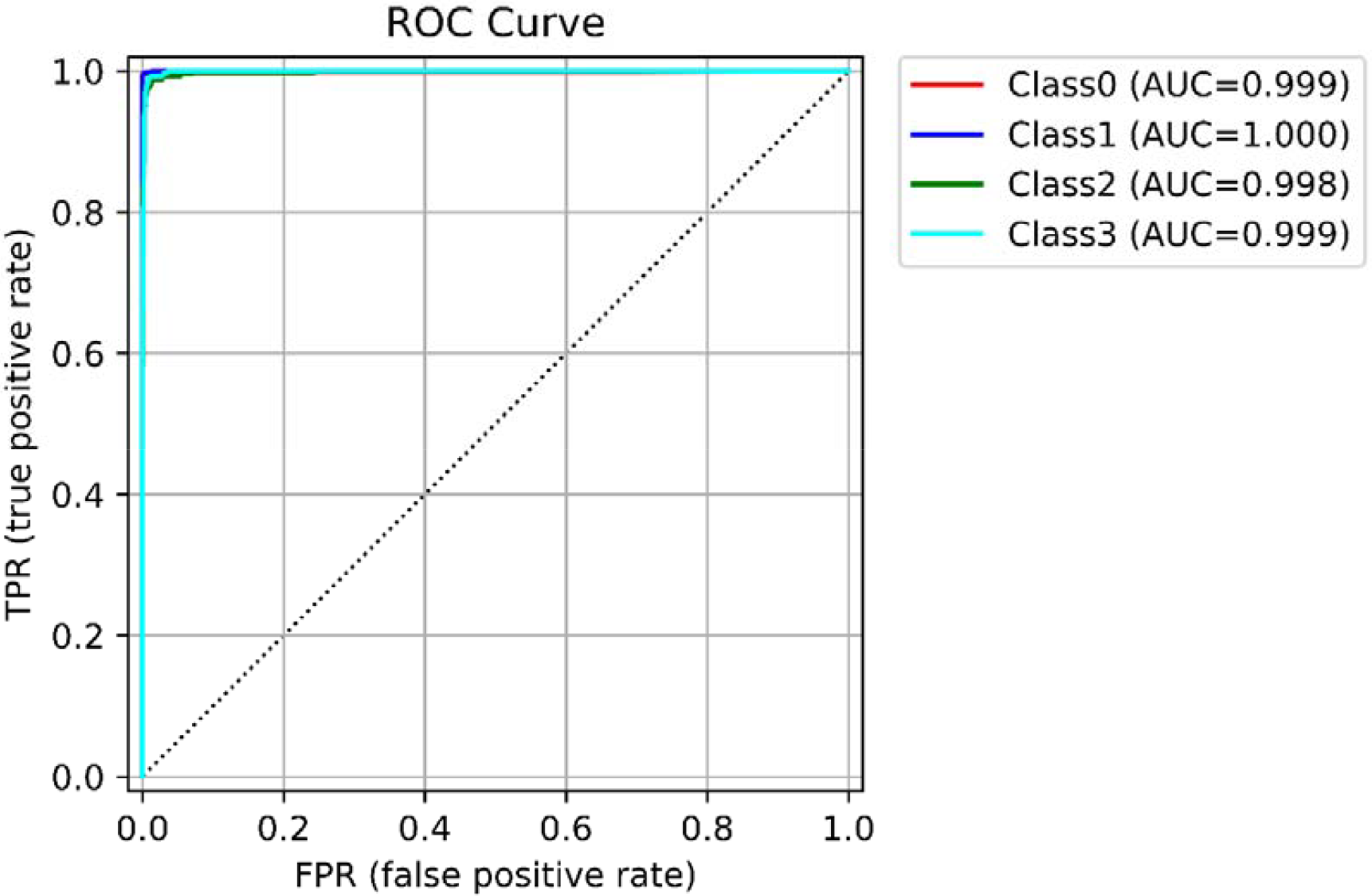
ROC curve showing excellent classifications. The legend at the top-right shows the calculated AUC for each class.

### Classification results – Ten-folds

Table 3 shows the performance of our proposed model for each fold, averaged over all four classes. The results are consistent among folds which shows the robustness and reproducibility of our model.

**Table 3.** The classification performance separated by folds.

| Fold | F1_score | sensitivity | specificity | precision | accuracy | AUC |
| --- | --- | --- | --- | --- | --- | --- |
| 0 | 0.9820 | 0.9831 | 0.9949 | 0.9810 | 0.9920 | 0.9988 |
| 1 | 0.9767 | 0.9821 | 0.9940 | 0.9734 | 0.9901 | 0.9998 |
| 2 | 0.9777 | 0.9799 | 0.9937 | 0.9759 | 0.9901 | 0.9973 |
| 3 | 0.9952 | 0.9940 | 0.9986 | 0.9964 | 0.9980 | 0.9999 |
| 4 | 0.9856 | 0.9846 | 0.9960 | 0.9869 | 0.9940 | 0.9998 |
| 5 | 0.9880 | 0.9892 | 0.9960 | 0.9870 | 0.9940 | 0.9995 |
| 6 | 0.9810 | 0.9812 | 0.9949 | 0.9811 | 0.9920 | 0.9975 |
| 7 | 0.9821 | 0.9808 | 0.9946 | 0.9835 | 0.9920 | 0.9997 |
| 8 | 0.9903 | 0.9881 | 0.9972 | 0.9930 | 0.9960 | 0.9999 |
| 9 | 0.9904 | 0.9904 | 0.9974 | 0.9904 | 0.9960 | 0.9998 |

Supplementary figure 2 shows the normalized confusion matrix for each fold, demonstrating consistent performance.

## Discussion

Identifying the contrast injection phase in contrast-enhanced CT images is a crucial step in using online or local databases for various machine learning or modelling tasks. Unfortunately, the DICOM headers, and naming in metadata among imaging centers is not standardized, which is a hurdle in developing models based on large datasets. We proposed a novel method based on DL-based segmentation developed in our group and a RF machine learning model to classify CT images. Our results were promising achieving an average AUC of 0.9991 and accuracy of 0.9936. Ten-fold data split with consistent performance over all folds demonstrated the robustness of our methodology on a fairly large database consisting of 2509 3D CT images. The overall error in our study is an accumulation of errors arising from the segmentation and machine learning classification tasks. However the whole pipeline is automated from scratch and showed excellent results. Our light-weight segmentation model with an average inference time of ∼8 seconds per image for all seven organs and less than one second for machine learning part offers an affordable and accurate methodology for handling large datasets. We attempted to reproduce what human logic follows to indicate the contrast media phase in a typical CT image, which makes our methodology completely explainable. After intra-venous injection of the contrast media, depending on the time passed, different combinations of enhancement in different organs are observed. First, severe enhancement in the descending aorta, then the rest of organs as shown in Figure 2 and Figure 3. We used the basic information from these important organs to predict the injection phase. To do so, the input to machine learning classifier is a very basic statistical measurement of the average, SD, and 10, 50, and 90 percentiles. Boruta feature selection selected all 35 features which was expected as we only calculated meaningful first-order features. Our aim was to make the procedure as simple and explainable as possible without using deep learning classifiers and without using higher-order radiomics/texture features. By breaking down the decision process into segmentation and classification steps, we tried to avoid using AI as a black box and follow the human logic to perform the task.

In terms of accuracy, our proposed model outperformed or is comparable with results reported in the published literature, with prediction of only 33/2509 cases (∼ <1.4%) in the wrong class. The accuracy was excellent for all four classes without a drop in performance in a single class. Our F1-score (0.9852) was better than the one reported by Zhou et al. [17] (0.977) and Dao et al. [19] (0.9209). The achieved accuracy (0.9936) was also higher than the one reported by Tang et al. [18] (0.93) and Muhamedrahimov et al. [20] (0.933).

Our study has a lot in common with what Reis et al. study [21]. The authors have almost followed the same steps, except that they used 48 radiomics features from each organ to predict the contrast media phase and used TotalSegmentator [33] trained models to generate organs masks. The number of features was lower in our study (five simple features), and we used our own segmentation model [23], which is much faster than TotalSegmentor [33] (ten seconds vs three minutes). In terms of accuracy, our model outperformed their reported results. The F1-scores (our study vs Ries et al. study) were 0.9978 vs 0.966, 0.9951 vs 0.789, 0.9667 vs 0.922, and 0.9813 vs 0.950 for the four classes consisting of non-contrast, arterial, venous, and delayed, respectively.

Because of using multiple organs information, our model may be robust for cases with pathological conditions, such as renal failure, which can affect deep learning-based image classification methodologies [15]. It should be mentioned that our segmentation model was inferred on each CT image at each phase and performed well on images from different contrast media injection phases. Due to the fact that our model inference is very fast (1 second per image/organ), generating the training dataset was not time consuming. Besides, our dataset contains patients referred to radiology department for severe pathologies in the liver and the segmentation module showed robust performance in those cases (one of these cases is shown in Figure 2).

Among the limitations of this study is that the adopted methodology depends on the accuracy of the segmentation model. However, if the segmentation model misses some parts of the aorta, liver, heart, or other organs, the average value and the simple features we selected and used are not significantly affected. We used the average value in the aorta and other organs for partly segmented aorta and the average difference was less than 5%.

## Conclusion

We developed a fast, accurate, reliable, and explainable methodology to classify the contrast media phases in CT images, which may be useful in data curation and annotation of large online datasets or local datasets with non-standard or no series description. This algorithm might help in better exploitation of shared datasets more effectively and help to gain better solutions for a variety of AI tasks by providing more reliable data tags and information about contrast media phases.

## Data Availability

The dataset is not publicly available.

## Acknowledgements

This work was supported by the Euratom research and training programme 2019-2020 Sinfonia project under grant agreement No 945196 and by the Distinguished Professor Program of Óbuda University, Budapest Hungary.

## Supplementary materials

Supplementary figure 1 compares the performance of different combinations of feature selection and models in classification task.

**Supplementary figure 1.**
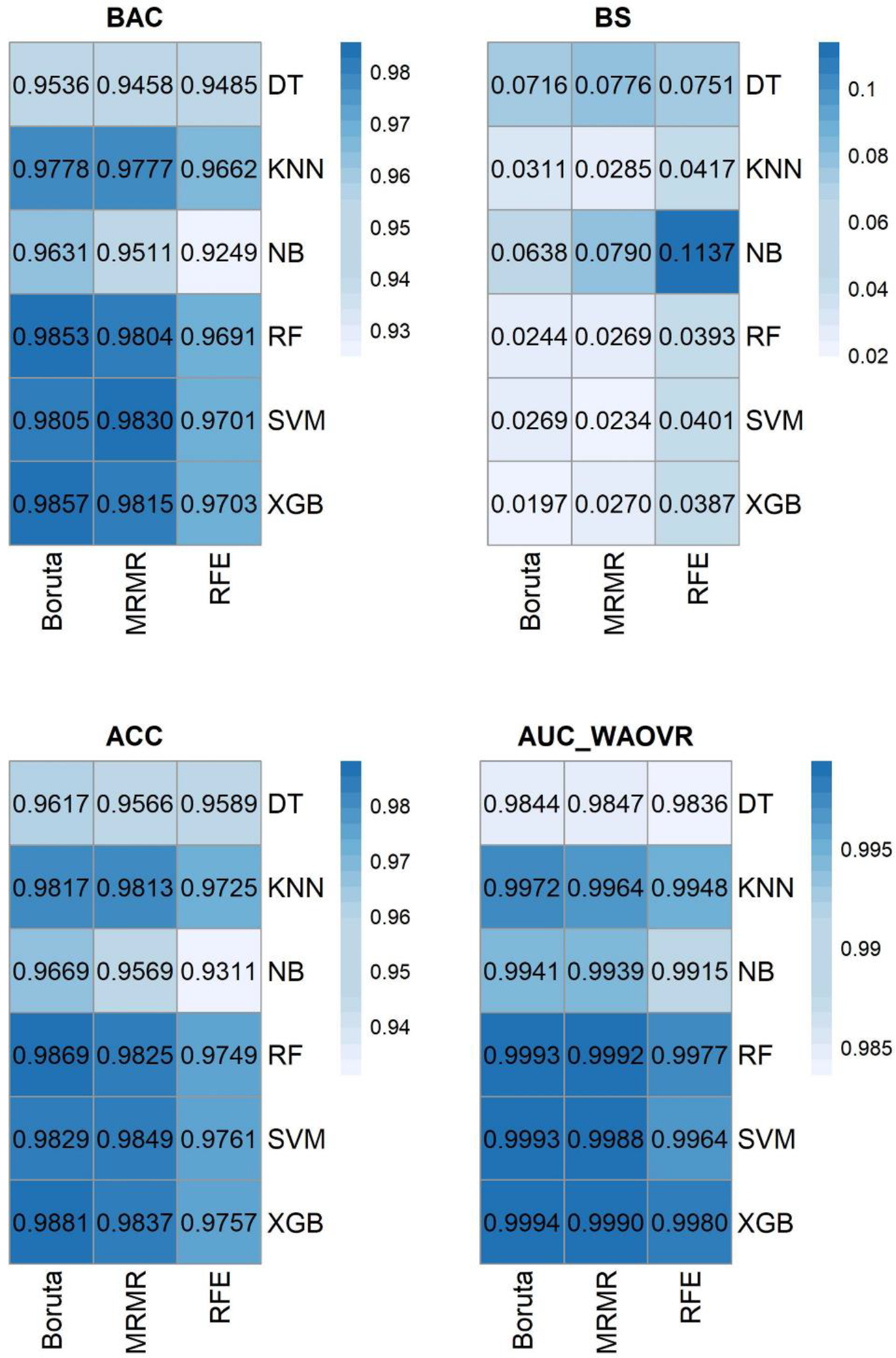
Comparison of the performance of models and feature selection, ACC: Accuracy, AUC_WAOVR: Weighted average one vs rest AUC, BS: Bier Score, BAC: Balanced accuracy.

Supplementary figure 2 shows the normalized confusion matrix of the best model (Boruta - RF) separately for each fold.

**Supplementary figure 2.**
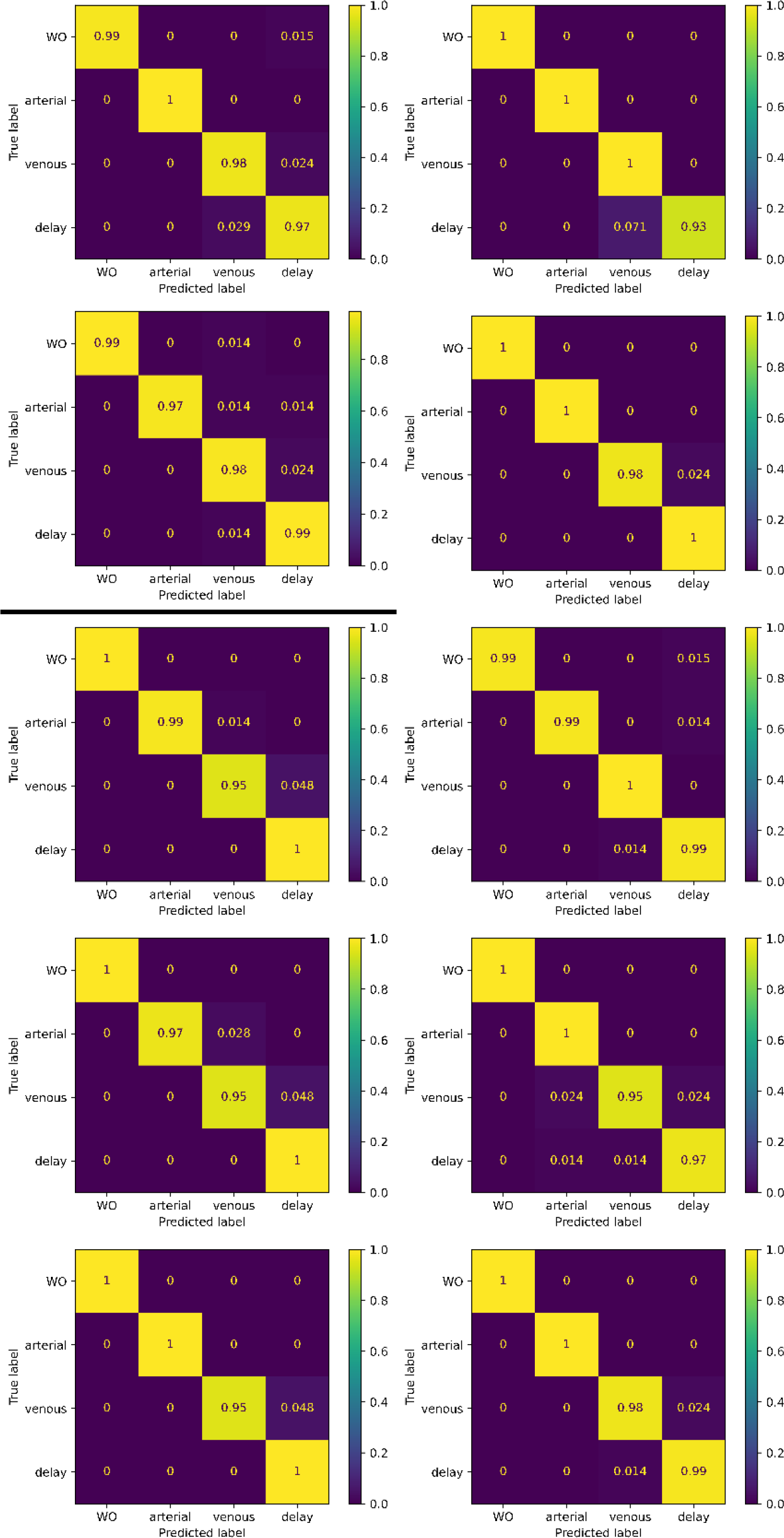
Normalized confusion matrices for all ten folds.

## Notes

### Competing Interest Statement

The authors have declared no competing interest.

### Author Declarations

The study was approved by the institutional ethics committee of HUG (CCER ID: 2017-00922) which allows us to process these images retrospectively.

